# From free text to SOFA score: automated reconstruction of sepsis severity from unstructured clinical notes

**DOI:** 10.64898/2025.12.17.25342509

**Authors:** Katherine Monsalve, Natalia Castano-Villegas, Elmer Escandón, José Zea, Laura Velásquez

**Affiliations:** Physician and Epidemiologist, AI Scientist, Arkangel AI, Bogotá, Colombia; Physician and Epidemiologist, CID Specialist – Research Leader, Arkangel AI, Bogotá, Colombia; Machine Learning Engineer, Arkangel AI, Bogotá, Colombia; Chief Executive Officer, Arkangel AI, Bogotá, Colombia; President, Arkangel AI, Bogotá, Colombia

**Keywords:** Sepsis, SOFAS scores, natural language processing, electronic health records, critical care

## Abstract

**Objective:** To evaluate the ability of a natural language processing system to automatically reconstruct the SOFA score from unstructured clinical notes in patients with sepsis and validate its applicability in intensive care units.

**Materials and methods:** Retrospective study in the MIMIC-III database that included 284 adults with sepsis. The SOFA calculated with structured data was compared with the SOFA reconstructed by free text extraction. Clinical rules were applied for calculation at 24 h and 48 h. Variable completeness, severity reclassification, and association with hospital mortality were evaluated using logistic regression.

**Results:** Automated extraction increased the availability of critical variables (respiratory 33% to 100%, vasopressor 12% to 41%). The reconstructed SOFA increased by 3 points at 24 hours, reclassifying patients with high severity (SOFA ≥ 6) from 17% to 48% and SOFA ≥ 10 from 5% to 22%. Reconstructed scores remained associated with mortality at 24 h (OR 1.16, 95% CI 1.09-1.24) and at 48 h (OR 1.23, 95% CI 1.15-1.31), comparable to that based on structured data (p < 0.001).

**Discussion:** Automatic reconstruction of the SOFA from free text recovers information missing from structured fields, reducing underestimation of severity.

**Conclusion:** NLP approaches supported by large language models provide a more complete and clinically consistent SOFA score in sepsis when structured data are insufficient.

## INTRODUCTION

Recent advances in large language models (LLMs) have substantially expanded the ability to process and structure clinical information contained in free-text narratives^1,2^. Within electronic health records (EHRs), clinical notes represent one of the richest sources of longitudinal and descriptive information, yet they remain among the most underutilized components for clinical analytics^3^.

A substantial proportion of clinically relevant information is embedded within thousands of pages of unstructured narrative documentation, rendering manual extraction labor intensive, error prone, and impractical at scale^4^. In this context, LLMs offer increasing potential to automate clinical entity extraction, reconstruct temporal trajectories of patient events, and derive complex variables directly from clinical text, thereby improving both clinical decision making and data-driven research^5,6^.

In critical care settings, these capabilities are particularly relevant. Sepsis remains one of the leading causes of hospital mortality and a major global public health challenge, accounting for approximately 11 million deaths annually nearly 20% of all global mortality^7^. Early identification and continuous monitoring of organ dysfunction are essential for prognosis and clinical decision making^8^.

The Sequential Organ Failure Assessment (SOFA) score is central to assessing disease severity, tracking clinical evolution, and guiding therapeutic decision making in sepsis^9^. However, its routine calculation is constrained by the fragmentation of key variables across structured fields and extensive unstructured clinical notes, which hinders the reproducibility and automation of clinical severity metrics^10^. Publicly available clinical datasets, such as MIMIC-III, contain much of the information needed to calculate SOFA, including vasopressor use, neurological assessments, ventilatory parameters, and laboratory results, and provide an ideal environment for evaluating this approach under real-world clinical documentation conditions.

Despite rapid advances in natural language processing (NLP) and large-scale language models applied to healthcare, a substantial gap persists between their theoretical potential and their effective deployment in critical care environments^11^. In particular, there is a lack of robust clinical validations demonstrating their ability to reconstruct complex physiological variables and enhance severity estimation in contexts where the volume, heterogeneity, and complexity of narrative documentation pose significant challenges^12^.

In this study, we evaluate the utility of Arkangel AI, a clinical natural language processing system designed to extract structured, clinically meaningful information from unstructured notes. For this work, Arkangel was specifically configured to reconstruct the SOFA score from narrative documentation, and its performance was compared against structured data available in MIMIC-III^13^ in the context of sepsis. Our objective is to assess its operational viability and clinical coherence, as well as its ability to process extensive ICU narratives under real-world conditions, providing evidence of its applicability in critical care settings.

## METHODS

### Study Design

A retrospective cohort study was conducted using electronic health records (EHRs) from adult patients admitted to the Intensive Care Unit (ICU) in the Medical Information Mart for Intensive Care III (MIMIC-III, version 1.4), a publicly available critical care database approved by the Institutional Review Boards of the Beth Israel Deaconess Medical Center and the Massachusetts Institute of Technology. Access to the dataset was obtained through completion of the required PhysioNet research certification.

### Population and Eligibility Criteria

We included all records corresponding to the first septic episode of patients hospitalized in the ICU. Cases were identified using ICD-9 codes in the structured *diagnoses_icd* table, including 995.91 (sepsis), 995.92 (severe sepsis), 785.52 (septic shock), and the 038.xx group corresponding to specific and unspecified septicemias. Only episodes with a total hospital stay of ≤ 30 days and a single eligible admission per patient were retained.

We excluded duplicate admissions, ICU stays < 24 hours, and records with inconsistencies that prevented linkage across the clinical files associated with the episode.

### Cohort Construction and Group Assignment

The cohort was constructed through a sequential filtering process described in Figure 1. From an initial set of 14,066 records with sepsis-related diagnoses, we identified all cases that met the eligibility criteria.

**Figure 1.**
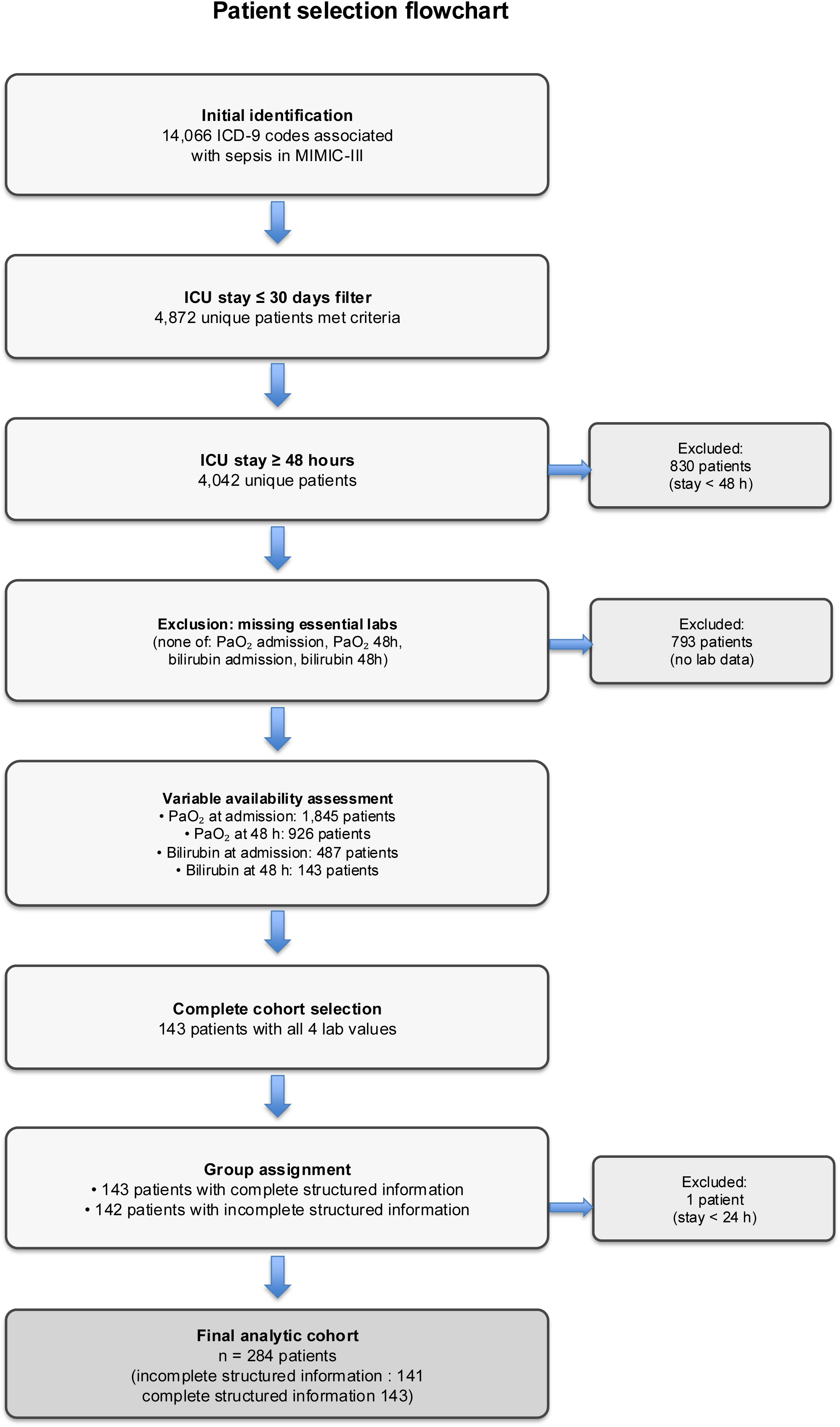
Study flow diagram

The structured-data cohort consisted of 143 patients with complete information, including availability of the minimum laboratory parameters required for SOFA calculation (PaO□/FiO□ratio and total bilirubin) at both ICU admission and 48 hours.

An additional 142 patients were randomly selected from the remaining eligible ICU admissions to form the cohort with incomplete structured information, which was appropriate for evaluating automated variable reconstruction from unstructured clinical text. One patient was excluded due to an ICU stay < 24 hours, resulting in 141 patients in this cohort. The final analytic cohort comprised 284 patients.

### Volume and characteristics of clinical text

The unstructured clinical notes associated with the 284 patients (143 with complete structured information and 141 with incomplete structured information) comprised approximately 8.7 million characters (≈ 1.45 million words), equivalent to roughly 2,900 pages of ICU documentation, including physician notes, nursing notes, ventilatory reports, and clinical orders. A detailed breakdown of narrative volume and processing time per patient is provided in Supplementary Table S1.

### Automated extraction procedure with Arkangel AI

Arkangel AI (previously referred to as Pandora in its initial version) is a modular artificial intelligence system that integrates two large language models with advanced natural language processing techniques and a clinical rule engine to automate the extraction of information from free text and reconstruct complex physiological variables^13^.

In this study, Arkangel processed all clinical notes directly, without manual preprocessing, and exhaustively extracted clinical conditions, vital signs, physiological parameters, and laboratory measurements explicitly documented in narrative text and embedded tables. Extraction was performed under a predefined set of constraints designed to ensure documentary fidelity, avoiding any form of clinical inference or reinterpretation of the recorded information (Supplementary Material 1).

All extracted elements were supported by verbatim textual excerpts from the source documents, ensuring traceability and clinical coherence. The resulting structured information was then organized into a relational database model and arranged in tables that enabled querying and aggregation using Structured Query Language (SQL).

From this structured representation, measurements corresponding to ICU admission and 48 hours were identified. In accordance with established SOFA clinical criteria, when more than one measurement was available within a given time window, score reconstruction was operationalized by selecting the lowest recorded values for PaO□, platelet count, and Glasgow Coma Scale; the highest values for bilirubin and creatinine; and the most severe cardiovascular state based on mean arterial pressure and vasopressor use.

### Internal validation of SOFA reconstruction

Internal validation involved confirming that the extracted values corresponded to explicit evidence within the clinical documentation, assessing temporal consistency across physiologically related variables, and performing a manual clinical review of a sample of ten cases to verify physiological plausibility and coherence with the documented clinical course.

### Structured and unstructured data sources

Structured data included laboratory results, vital signs, blood gas measurements, ventilatory parameters, administered medications, urine output, and Glasgow Coma Scale when available. These data were obtained from the PATIENTS, LABEVENTS, CHARTEVENTS, INPUTEVENTS, and OUTPUTEVENTS tables of MIMIC-III.

Unstructured data consisted of all physician notes, nursing notes, ventilatory reports, and narrative documents associated with each clinical episode. These sources were processed using Arkangel AI to extract the variables required for SOFA calculation, including PaO□, FiO□, the PaO□/FiO□ ratio, total bilirubin, platelet count, creatinine, urine output, Glasgow Coma Scale, vasopressor use, additional ventilatory parameters, and SOFA subsystem scores. All extracted values were aligned within the 0–24-hour and 48-hour (± 6 hours) windows.

To further support extraction fidelity, a manual review of 10 randomly selected patients was performed (Supplementary Table S2), showing high concordance between structured values, narrative documentation, and Arkangel outputs.

For each patient, two matrices were generated: a pre-Arkangel structured matrix derived exclusively from tabulated variables available in MIMIC-III, and a post-Arkangel extraction matrix incorporating automatically extracted values from unstructured text. These matrices were compared to evaluate recovery of missing data, completeness by the SOFA component, variation in global SOFA scores, and changes in severity categories defined by SOFA thresholds (≥2, ≥6, and ≥10).

### Operational performance

Operational performance was assessed through the processing time per patient, the total time required to process the cohort, and the volume of text analyzed per case. Arkangel processed each patient in 1–15 minutes depending on note size (Small <10,000 characters; Very Large >200,000 characters), with a total parallel processing time of approximately two hours for the entire cohort, compared with the hours or days typically required for manual review.

### Study variables

The primary outcome was the completeness of SOFA score criteria before and after automated processing with Arkangel AI. This outcome was evaluated across all SOFA domains using the clinical and laboratory variables required for score construction, including PaO□, FiO□, PaO□/FiO□ ratio, platelet count, total bilirubin, creatinine, mean arterial pressure, urine output, and Glasgow Coma Scale. These variables enabled derivation of individual SOFA subsystem scores and assessment of physiological coherence in the reconstructed SOFA.

Secondary outcomes included changes in total SOFA score between ICU admission and 48 hours, transitions across clinically relevant severity categories (SOFA ≥2, ≥6, and ≥10), and the impact of Arkangel AI–based extraction on predictive performance for 30-day mortality. Additional descriptive variables included age, admission classification (sepsis versus septic shock), vasopressor use, and need for intubation within the first 24 hours.

### Statistical methods

Continuous variables were summarized as medians with interquartile ranges due to the non-parametric distribution of scores. Categorical variables were expressed as absolute and relative frequencies, with group comparisons (complete structured information vs incomplete structured information) performed using chi-squared or Fisher’s exact test as appropriate. Pre–post comparisons at 24 and 48 hours were assessed using the Wilcoxon signed-rank test, and differences in SOFA change between groups were evaluated with the Mann–Whitney test. Logistic regression models were fitted to predict 30-day mortality using pre- and post-extraction SOFA scores, reporting odds ratios and 95% confidence intervals. Predictive performance was evaluated using ROC curves, area under the curve (AUC) with bootstrap confidence intervals, and calibration using the Brier score. Net reclassification improvement (NRI) and integrated discrimination improvement (IDI) were calculated as supplementary analyses. All analyses were conducted in Python 3.9.6.

### Ethical considerations

This study used exclusively secondary data that were fully de-identified and publicly available. In accordance with international regulations governing the use of anonymized data (US Common Rule, 45 CFR 46) and Colombian regulations (Resolution 8430 of 1993), this type of analysis is considered minimal-risk research, is exempt from institutional ethics committee review, and does not require informed consent^14,15^. Although the database already ensured anonymization, all data handling and analyses were conducted following general ethical principles consistent with the Belmont Report and the Declaration of Helsinki, ensuring responsible and respectful use of clinical information^16,17^.

## RESULTS

At baseline, prior to Arkangel AI processing and using only structured data, both cohorts showed no major differences in sex, initial clinical classification, or vasopressor use within the first 24 hours (all p > 0.05). The complete structured-data cohort exhibited a higher frequency of early intubation (44.8% vs. 31.2%; p = 0.026). Differences in several continuous parameters derived from structured data were observed, including platelet count (227 vs. 195 ×10³/μL; p = 0.001), bilirubin (1.1 vs. 0.8 mg/dL; p = 0.006), Glasgow Coma Scale (5 vs. 10 points; p < 0.001), and creatinine (1.3 vs. 1.0 mg/dL; p = 0.021). Consistently, SOFA subscores based on structured data were higher in the coagulation, hepatic, and renal domains (all p < 0.05), resulting in a slightly lower total SOFA score at 24 hours in the incomplete structured-data cohort (6.0 vs. 9.6; p < 0.001) (Table 1).

**Table 1.**
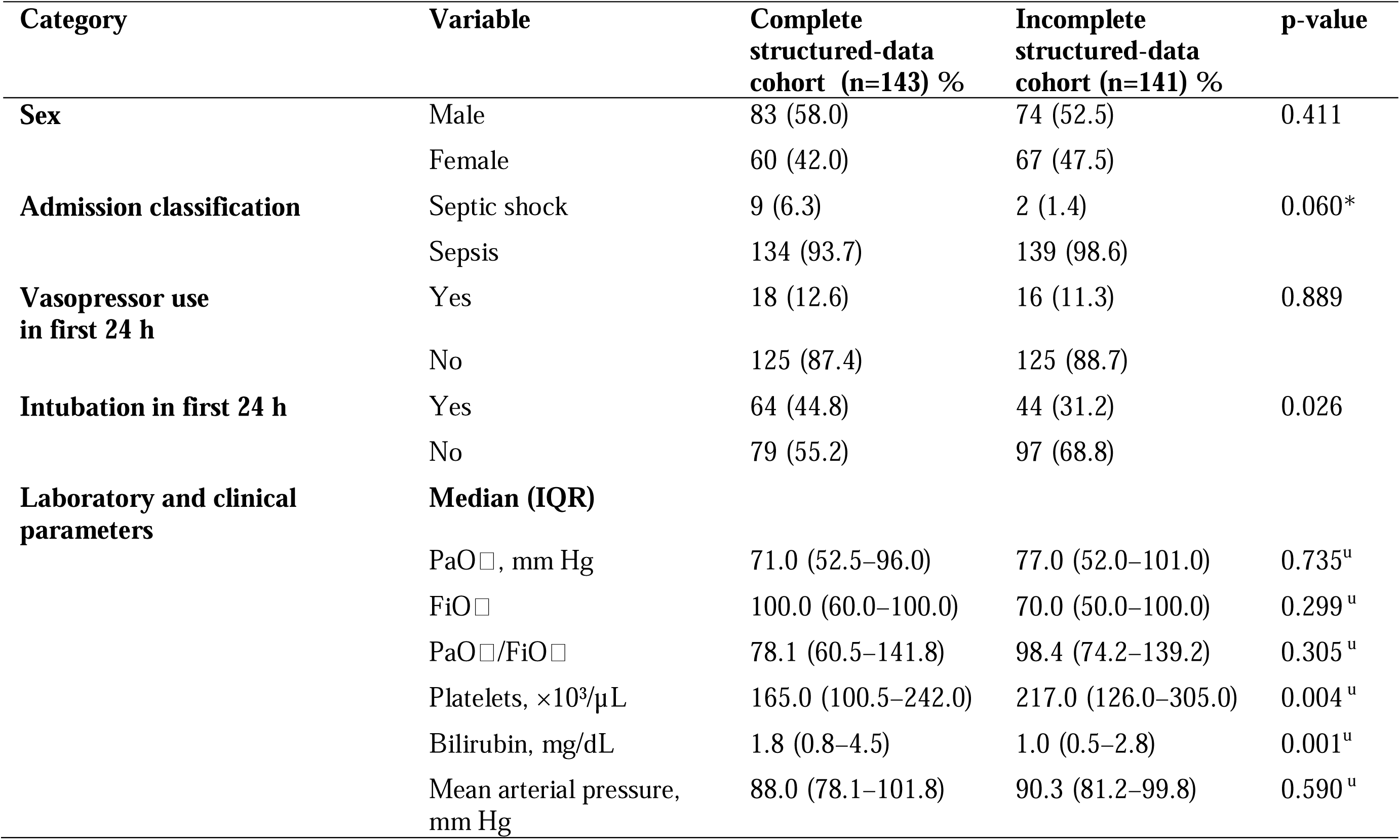
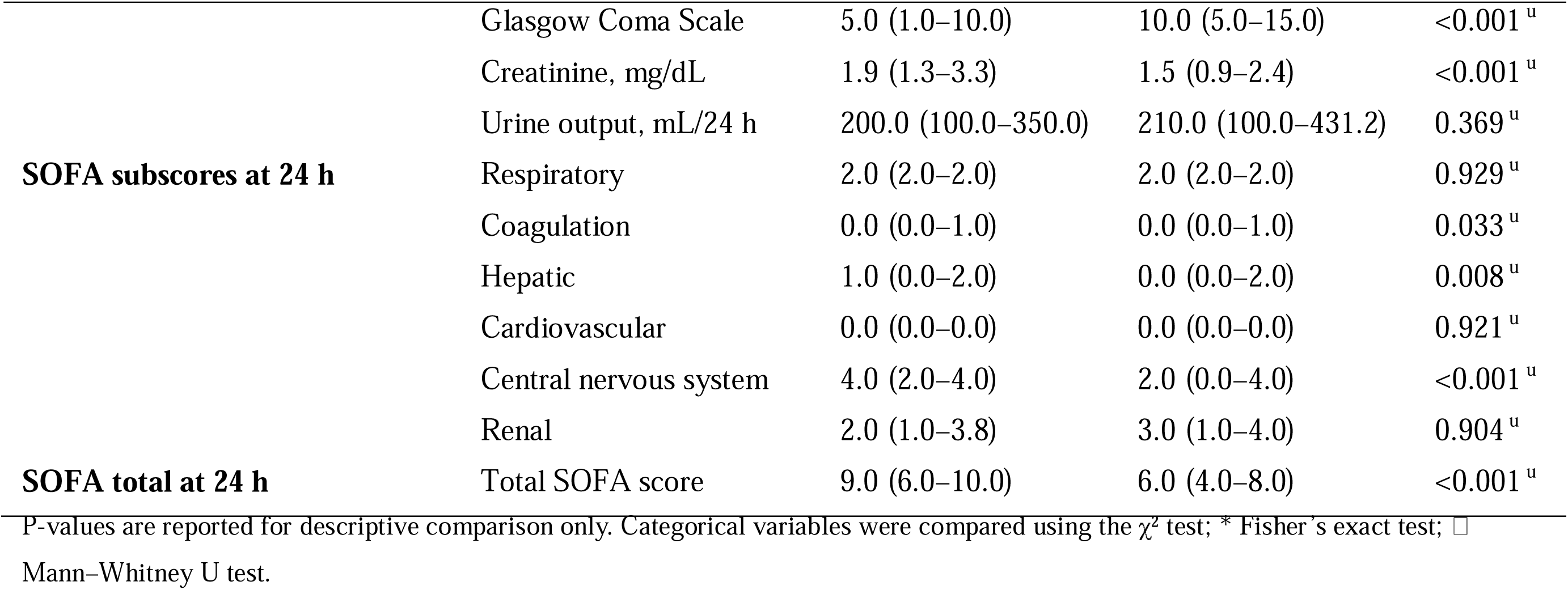
Baseline Characteristics of the Study Cohorts (Structured Data Only)

Data availability required for SOFA calculation showed substantial improvements after extraction, particularly in respiratory variables. At 24 hours, FiO□ availability increased from 33.8% to 100% and PaO□ from 92.6% to 100%, enabling the PaO□/FiO□ ratio to rise from 33.1% to 100%. Increases were also observed in mean arterial pressure (77.8% to 98.2%), vasopressor documentation (12.0% to 41.2%), total bilirubin (76.4% to 77.1%), and urine output (52.5% to 58.1%). In contrast, slight decreases were noted in the availability of platelet count (99.6% to 99.3%) and creatinine (99.6% to 98.2%).

A similar pattern was observed at 48 hours, with notable increases in FiO□ (36.3% to 100%), PaO□/FiO□ ratio (33.1% to 94%), and mean arterial pressure (62.0% to 87.3%), accompanied by small negative variations in creatinine, Glasgow Coma Scale, and platelet count. (Table 2)

**Table 2.** Data Availability for SOFA-Related Variables Before and After Extraction (24 h and 48 h)

| Variable | SOFA System | Pre 24 h<br>n=284<br>(%) | Post 24 h<br>n=284<br>(%) | Δ 24 h (%) | Pre 48 h<br>n=284<br>(%) | Post 48 h<br>n=284<br>(%) | Δ 48 h (%) |
| --- | --- | --- | --- | --- | --- | --- | --- |
| FiO<sub>2</sub> | Respiratory | 33.8 | 100 | 66.2 | 36.3 | 100 | 63.7 |
| PaO<sub>2</sub> | Respiratory | 92.6 | 100 | 7.4 | 81.7 | 94 | 12.3 |
| PaO<sub>2</sub>/FiO<sub>2</sub> | Respiratory | 33.1 | 100 | 66.9 | 33.1 | 94 | 60.9 |
| Platelets | Coagulation | 99.6 | 99.3 | −0.4 | 98.6 | 98.9 | 0.4 |
| Total bilirubin | Hepatic | 76.4 | 77.1 | 0.7 | 63.0 | 69.4 | 6.4 |
| Mean arterial pressure (MAP) | Cardiovascular | 77.8 | 98.2 | 20.4 | 62.0 | 87.3 | 25.4 |
| Vasopressors | Cardiovascular | 12.0 | 41.2 | 29.2 | 11.3 | 27.8 | 16.5 |
| Glasgow Coma Scale (GCS) | CNS | 98.2 | 98.2 | 0 | 97.7 | 97.9 | 0.2 |
| Creatinine | Renal | 99.6 | 98.2 | -1.4 | 98.6 | 97.2 | -1.4 |
| Urine output (mL/24 h) | Renal | 52.5 | 58.1 | 5.6 | 51.4 | 56.0 | 4.6 |

Δ: difference between post-extraction and pre-extraction availability.

The SOFA score increased after extraction in both groups and across both time windows. At 24 hours, the median SOFA score increased from 9.0 to 12.0 in the complete structured-data cohort and from 6.0 to 9.0 in the incomplete structured-data cohort (p < 0.001 for both comparisons). At 48 hours, SOFA increased from 9.0 to 11.0 in the complete structured-data cohort and from 6.0 to 7.0 in the incomplete structured-data cohort (p < 0.001 for both comparisons). The absolute change (ΔSOFA) was 3 points in the complete structured-data cohort at both 24 and 48 hours, and 3 and 2 points, respectively, in the incomplete structured-data cohort.

Reclassification analyses showed that, after extraction, a higher proportion of patients in each group met established SOFA severity thresholds. At 24 hours, the proportion of patients with SOFA ≥6 increased from 83.9% to 97.9% in the complete structured-data cohort and from 61.7% to 87.9% in the incomplete structured-data cohort, while SOFA ≥10 increased from 38.5% to 74.8% and from 17.0% to 48.2%, respectively. Similar patterns were observed at 48 hours. (Table 3).

**Table 3.** Pre- and post-extraction SOFA scores.

| Group | Time | SOFA pre, median (IQR) | SOFA post, median (IQR) | ΔSOFA, median (IQR) | p-Wilcoxon |
| --- | --- | --- | --- | --- | --- |
| complete structured-data cohort | 24 h | 9.0 (6.0–10.0) | 12.0 (9.5–14.5) | 3.0 (2.0–4.5) | <0.001 |
| Incomplete structured-data cohort | 24 h | 6.0 (4.0–8.0) | 9.0 (7.0–12.0) | 3.0 (2.0–4.0) | <0.001 |
| complete structured-data cohort | 48 h | 9.0 (7.0–11.0) | 11.0 (9.0–15.0) | 3.0 (1.0–4.0) | <0.001 |
| Incomplete structured-data cohort | 48 h | 6.0 (4.0–9.0) | 7.0 (6.0–11.0) | 2.0 (0.0–3.0) | <0.001 |

**Table 4.**
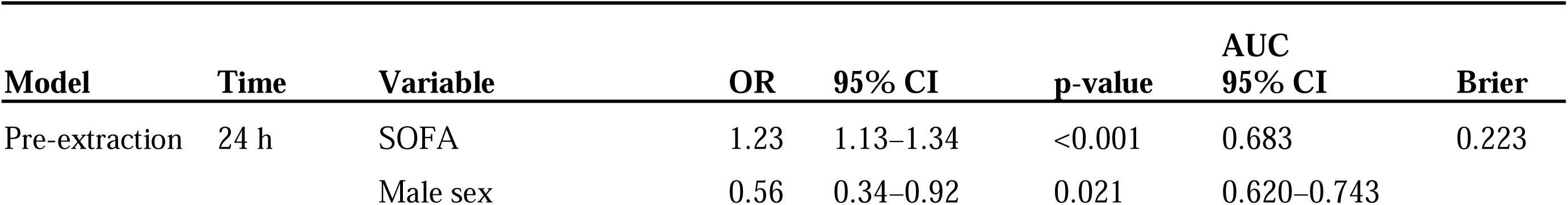
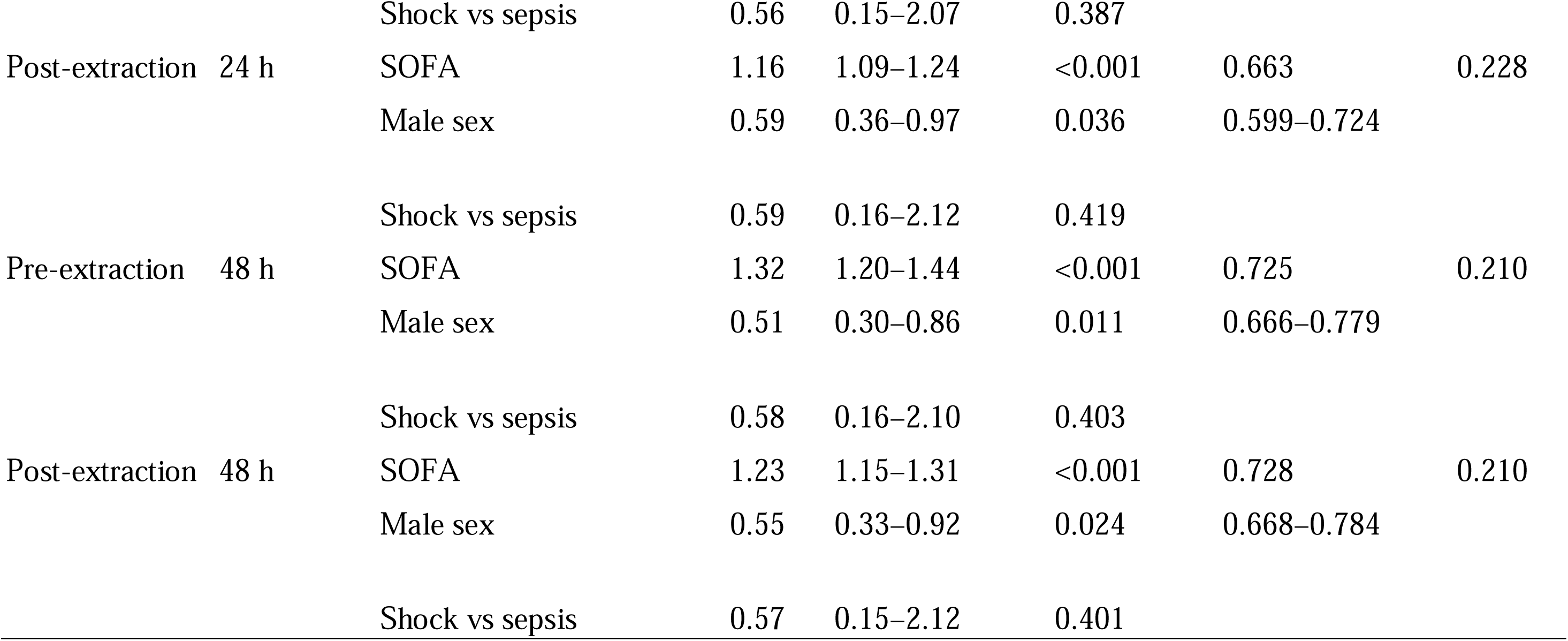
Logistic Regression Models Predicting In-Hospital Mortality Using SOFA Scores Before and After Data extraction.

To further visualize these changes, Figure 2 shows the distribution of ΔSOFA (post – pre) at 24 h and 48 h in both groups. Boxplots illustrate the median, interquartile range, and overall distribution of ΔSOFA within each cohort. Within-group comparisons were evaluated using the Wilcoxon signed-rank test (all p < 0.001), and between-group differences in ΔSOFA were assessed using the Mann–Whitney U test (24 h: p = 0.007; 48 h: p = 0.001). Figure 2.

**Figure 2.**
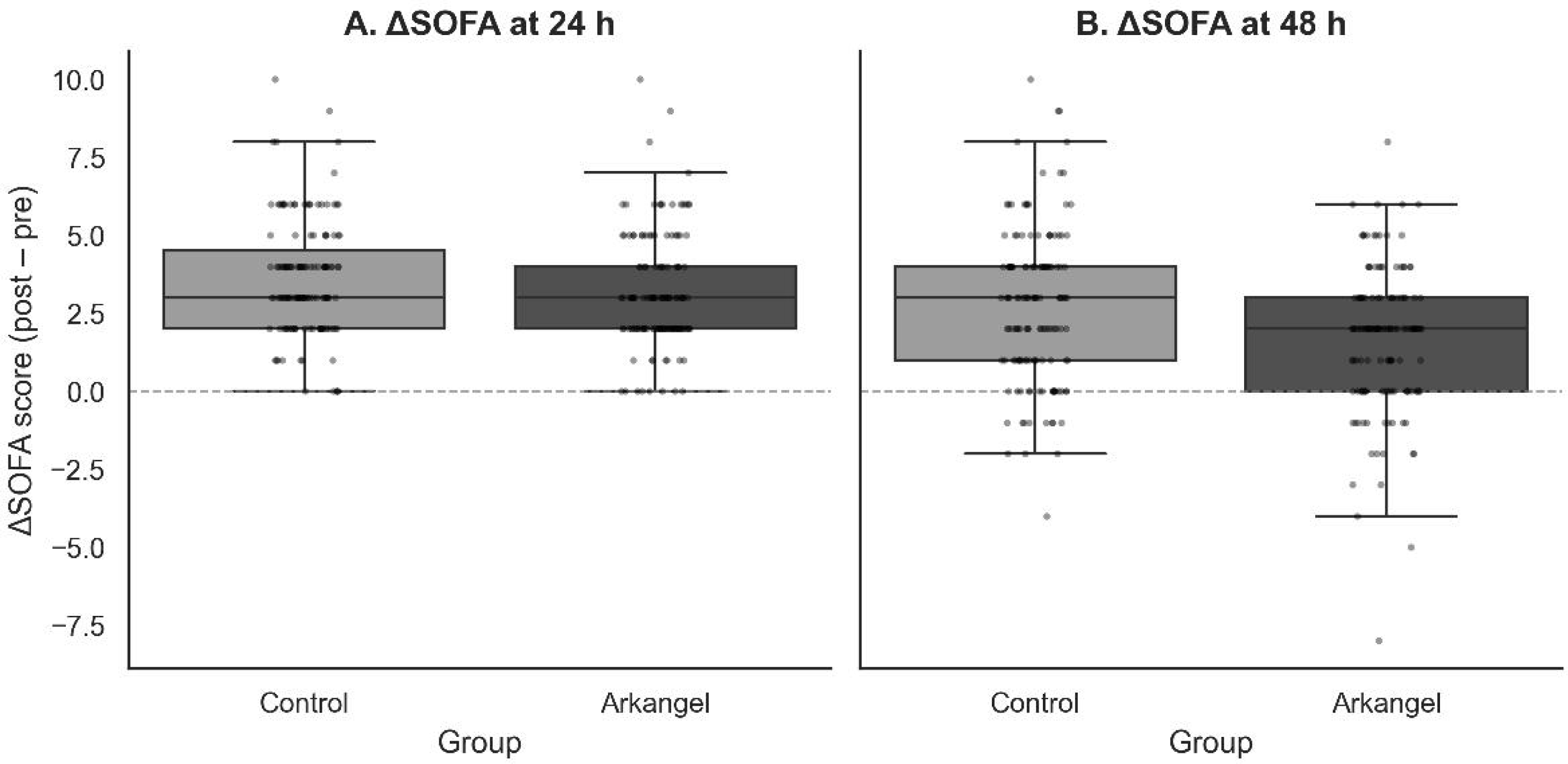
ΔSOFA scores at 24 h and 48 h in the complete structured-data cohort and Incomplete structured-data cohort

Total SOFA scores increased significantly after Arkangel extraction at both 24 and 48 hours in both groups (all p < 0.001). Figure 3 shows the redistribution of patients across established SOFA severity thresholds (≥2, ≥6, ≥10), with a higher proportion of patients meeting each threshold after extraction compared with pre-extraction values

**Figure 3.**
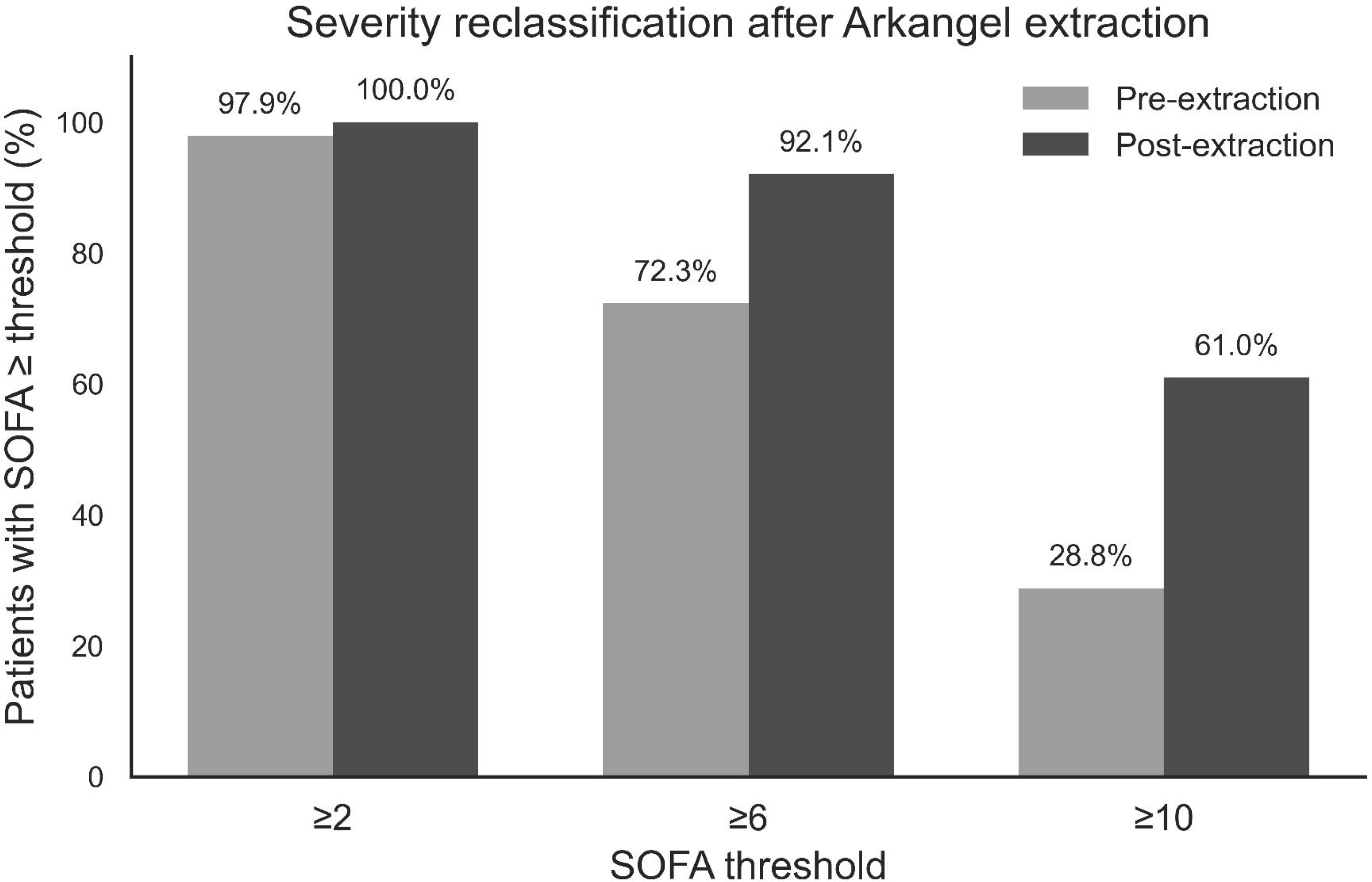
Severity reclassification (≥2, ≥6, ≥10)

To assess whether SOFA scores reconstructed after Arkangel AI processing retained their predictive power for mortality, a logistic regression model was estimated. At 24 hours, the pre-extraction SOFA score showed an odds ratio (OR) of 1.23 (95% CI: 1.13–1.34), while the post-extraction SOFA score showed an OR of 1.16 (95% CI: 1.09–1.24), with both associations reaching statistical significance (p < 0.001). Consistent results were observed at 48 hours (pre-extraction SOFA: OR 1.32, 95% CI: 1.20–1.44; post-extraction SOFA: OR 1.23, 95% CI: 1.15–1.31).

Male sex was associated with lower odds of mortality, with modest effect sizes and variable statistical significance, whereas initial clinical category (septic shock versus sepsis) was not significantly associated with mortality.

Performance metrics indicated acceptable discrimination, with AUC values ranging from 0.663 to 0.728 and Brier scores between 0.210 and 0.228, without meaningful differences between pre- and post-extraction estimates.

Receiver operating characteristic (ROC) curves were generated to compare mortality prediction performance using SOFA scores derived from structured data only versus SOFA scores reconstructed after automated extraction. Figure 4 shows ROC curves at 24 hours (panels A and B) and 48 hours (panels C and D). At 24 hours, the AUC values were 0.683 for structured-data SOFA and 0.663 after extraction; at 48 hours, AUC values were 0.725 and 0.728, respectively. Discriminative performance was comparable at both time points.

**Figure 4.**
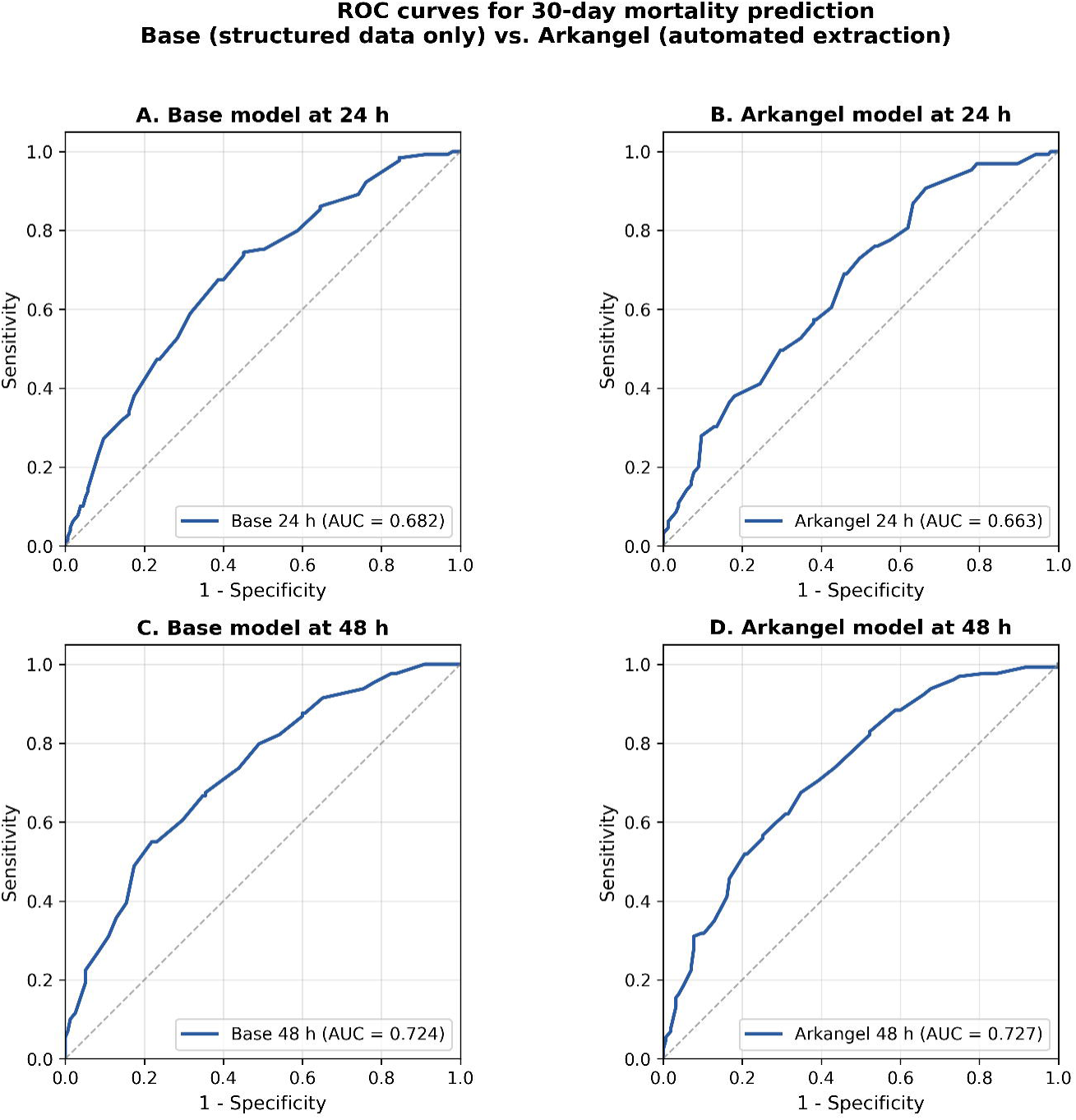
ROC curves for SOFA-based mortality prediction before and after automated extraction. Panel A: Pre-extraction (structured data only) at 24 h. Panel B: Post-extraction at 24 h. Panel C: Pre-extraction (structured data only) at 48 h. Panel D: Post-extraction at 48 h.

Additional reclassification metrics (NRI and IDI) comparing Arkangel and Base models are provided in Supplementary Table S3.

## DISCUSSION

This study demonstrates that the natural language processing (NLP) system Arkangel can reconstruct the SOFA score from unstructured clinical notes in a real-world critical care dataset. The analyzed cohort included 284 patients whose clinical free-text notes comprised approximately 8.7 million characters, equivalent to nearly 2,900 pages of clinical text a volume that makes systematic manual extraction virtually infeasible and reflects the operational challenge health systems face when transforming dispersed documentation into analytically usable data^1,18^.

One of the central findings was the asymmetry in the initial availability of structured data between groups. The complete structured-data cohort, which had the minimum laboratory data required for traditional SOFA calculation, represented only 1% of the 14,066 initial records with sepsis-related diagnoses. This highlights the limitations of structured-data availability for critical physiologic variables in clinical charts and underscores the gap between what is documented in routine clinical practice and what is readily available for computational analytics^19^. The incomplete structured-data cohort, in contrast, included patients with incomplete structured information, reproducing a more representative ICU scenario and allowing the evaluation of the model’s performance under realistic conditions of fragmented information.

Although both groups showed comparable baseline characteristics in sex (p = 0.411), initial clinical sepsis classification (p = 0.060), and vasopressor use within the first 24 hours (p = 0.889), the complete structured-data cohort exhibited a higher frequency of early intubation (44.8% vs. 31.2%; p = 0.026), which reflect differences in baseline clinical severity and the need for early respiratory support.

Among continuous variables derived from structured data, several differences were observed between cohorts, including lower Glasgow Coma Scale scores and higher creatinine and bilirubin levels in the cohort with complete structured information, as well as higher platelet counts in the cohort with incomplete structured information. These contrasts resulted in a difference in total SOFA score at 24 hours (9.0 vs. 6.0 points).

These differences should be interpreted with caution, as the cohorts were defined based on the availability of structured information rather than clinical or physiological criteria. In this context, the observed variation in SOFA subscores across domains and in total SOFA score at 24 hours is consistent with heterogeneity in data completeness at ICU admission, rather than with underlying differences in disease severity. Previous studies have shown that, while NLP-based approaches can substantially enhance data completeness, their performance is inherently conditioned by documentation practices, temporal ambiguity, and variability in narrative structure, underscoring the need for clinically grounded validation in real-world settings^20^.

Arkangel-based extraction produced substantial improvements in the availability of data required for SOFA calculation. At 24 hours, FiOD availability increased by 66.2%, while PaOD increased by 7.4%, enabling a 66.9% increase in the availability of the PaOD/FiOD ratio. Mean arterial pressure availability rose from 77.8% to 98.2%, and documentation of vasopressor use increased by 29.2%. Although slight reductions were observed for creatinine (from 99.6% to 98.2%) and platelet count (from 99.6% to 99.3%), these changes were minimal.

At 48 hours, the same pattern was replicated, with increases in PaOD/FiOD availability of 60.9% and mean arterial pressure availability of 25%. These findings indicate that Arkangel successfully retrieves critical information dispersed across multiple clinical notes, enabling complete SOFA reconstruction even when structured data are insufficient.

A small proportion of missing values persisted after automated extraction, mainly in platelet count and creatinine. This behavior corresponds to a deliberate design decision, in which Arkangel prioritizes data fidelity over forced completeness and therefore refrains from imputing values when multiple conflicting measurements exist without sufficient temporal resolution. This conservative strategy is consistent with current recommendations in clinical AI, which emphasize that incorporating ambiguous or conflicting data can introduce bias and undermine model reliability. By excluding uncertain measurements, Arkangel preserves the physiological coherence of reconstructed severity scores, even at the expense of minimal completeness^21,22^.

This mechanism also reflects the inherent complexity of standardizing physiological measurements documented in free-text clinical notes, where a single variable may appear as “platelets,” “Plt,” “Plq,” or even as partial counts. A technical strength of the system is its ability to recognize these variants and correctly map them to the appropriate clinical entity; however, when multiple expressions coexist without sufficient temporal clarity, Arkangel prioritizes accuracy and discards the imputation to prevent downstream errors in SOFA calculation. Although this strategy reduces post-extraction completeness by less than 2%, it improves the reliability on the retrieved material and underscores the importance of integrating structured and narrative data in real-world settings. This approach is also consistent with contemporary principles of uncertainty management in clinical AI^22^.

SOFA scores increased consistently after automated extraction in both cohorts. At 24 hours, SOFA increased from 9.0 to 12.0 in the complete structured-data cohort and from 6.0 to 9.0 in the incomplete structured-data cohort; at 48 hours, from 9.0 to 11.0 and from 6.0 to 7.0, respectively. The absolute change was 3 points in the complete structured-data cohort at both times and 3 and 2 points in the incomplete structured-data cohort.

Reclassification analyses showed a increase in the proportion of patients reaching clinically relevant severity thresholds. At 24 hours, the proportion with SOFA ≥6 increased from 83.9% to 97.9% in the complete structured-data cohort and from 61.7% to 87.9% in the incomplete structured-data cohort, while SOFA ≥10 increased from 38.5% to 74.8% and from 17.0% to 48.2%, respectively. These changes indicate that automated extraction enables a more complete and accurate representation of organ dysfunction by recovering clinically relevant information that is present in the electronic health record but not captured in structured fields.

From an operational standpoint, Arkangel processed the entire cohort in approximately two hours, with per-patient processing times ranging from one to fifteen minutes—contrasting with the days (or months) that would be required to review thousands of pages of clinical notes manually. This performance demonstrates the feasibility of large-scale NLP implementation and suggests the potential to generate physiological scores with minimal latency in retrospective studies and, eventually, in routine clinical workflows ^1^. Although artificial intelligence has demonstrated benefits across diverse clinical settings, most applications focus on therapeutic support or digital monitoring and do not address the recovery of complex physiological variables from free-text clinical notes^23,24^. In this context, Arkangel represents a shift towards AI systems that integrate directly with the realities of hospital documentation.

Physiological coherence was also preserved when evaluating predictive performance: both the pre-extraction and post-extraction SOFA scores were significantly associated with in-hospital mortality (ORs ranging from 1.16 to 1.32), and AUC values ranged from 0.663 to 0.728, with no evidence of degradation in discriminative ability after reconstruction with Arkangel. This stability suggests that free-text-based extraction not only improves score completeness but also preserves its clinical validity, reinforcing the utility of the reconstructed SOFA score for prognostic analyses and potential applications in clinical decision support.

Notwithstanding, comparisons between cohorts are conditioned by differences in structured-data completeness and should therefore not be interpreted as reflecting true similarities or differences in underlying clinical severity. The quality of the reconstruction depends on the accuracy, temporal consistency, and level of detail with which physiological variables are documented in the narrative, factors that show substantial variability within MIMIC-III^25^.

The absence of an exhaustive gold standard for all 284 cases limits the ability to estimate the system’s sensitivity and specificity, and the retrospective design prevents direct evaluation of how Arkangel might influence real-time clinical practice. Although the data originate from English-language notes, ICU documentation structure is relatively standardized, and Arkangel has been developed to operate across widely used clinical languages (English, Spanish, Portuguese, and French), supporting conceptual transferability. Nevertheless, variation in narrative styles, local documentation conventions, and record granularity may influence system performance, underscoring the need for evaluation in additional clinical and linguistic contexts^26^.

Despite these limitations, the findings suggest that natural language processing systems can reduce critical gaps in the availability of physiological data for the analysis of sepsis and other high-complexity syndromes. Arkangel’s ability to recover longitudinal information dispersed across multiple notes, reconstruct multicomponent physiological scores, and process large volumes of text within minutes indicates that such tools can complement and in some scenarios extend the exclusive reliance on structured data. This opens opportunities to improve the quality of clinical analytics, strengthen longitudinal studies, enable automated monitoring systems, and enhance decision-making in the ICU. Future research should evaluate its integration with early warning systems, its prospective performance in real-world clinical workflows, and its applicability across diverse and multilingual healthcare settings.

## CONCLUSION

Arkangel demonstrated that it is possible to reconstruct the SOFA score from unstructured clinical text, overcoming the historical dependence on structured, tabulated data and enabling a more precise characterization of sepsis trajectories in scenarios where such reconstruction was previously unfeasible. This study also shows that clinical narrative can be transformed into a structured source of physiological information suitable for integration into clinical and computational analyses.

The system’s ability to process thousands of pages within minutes substantially expands the possibilities in retrospective research and in the monitoring of critically ill patients, opening the possibility for automated extraction and timely analysis to support decision-making in complex clinical environments.

## Data Availability

The data used in this study are derived from the MIMIC-III critical care database, which is publicly available to qualified researchers upon completion of the required data use agreement and training. No new data were generated for this study. All analyses were conducted using de-identified data in accordance with the database governance policies.

https://physionet.org/content/mimiciii/1.4/

## ACKNOWLEDGMENTS

We thank Luis David Roa for his contribution to the production of the video abstract associated with this manuscript. We also acknowledge the MIT Laboratory for Computational Physiology and the Beth Israel Deaconess Medical Center for providing access to the MIMIC-III database.

